# Cross-ancestry performance of Parkinson’s disease polygenic risk scores in admixed Latin American populations

**DOI:** 10.64898/2026.03.02.26347226

**Authors:** Victor Flores-Ocampo, Paula Reyes-Pérez, Natalia S. Ogonowski, Grecia Sevilla-Parra, Santiago Díaz-Torres, Thiago P. Leal, Emily Waldo, Alejandra E. Ruíz-Contreras, Sarael Alcauter, Paola Arguello-Pascualli, the Global Parkinson’s Genetics Program (GP2), Ignacio F. Mata, Miguel E. Rentería, Alejandra Medina-Rivera, Jessica K. Dennis

## Abstract

Parkinson’s disease (PD) is a disabling neurodegenerative disorder with a substantial heritable component. Despite major advances in genome-wide association studies (GWAS), polygenic risk scores (PRS) show reduced predictive performance outside European populations, limiting equitable translation. Latin American populations represent a particularly difficult case because of their characteristic three-way admixture. We evaluated the cross-ancestry transferability of PD PRS in 1,872 PD cases and 1,443 controls of Latin American ancestry using data from the Global Parkinson’s Genetics Program (GP2). PRS were constructed using summary statistics from a large European-ancestry GWAS, a moderately sized mixed-ancestry GWAS meta-analysis, and a small ancestry-matched Latin American GWAS. We benchmarked two single-ancestry approaches (PRSice-2 and SBayesRC) against two multi-ancestry methods (PRS-CSx and BridgePRS) that explicitly model cross-population genetic architecture. Across all performance metrics, SBayesRC performed best. PRS derived from large European GWAS achieved the highest effect size (odds ratio = 2.02; pseudo-R² = 0.031) while PRS derived from mixed ancestry GWAS meta-analysis yielded the highest discriminative ability (AUC=0.67). Our findings demonstrate that, under current sample size imbalances, well-powered European discovery GWAS outperform ancestry-matched but underpowered datasets in three-way admixed populations. Incorporating functional annotations, as implemented in SBayesRC, improves portability across ancestries. However, the full potential of multi-ancestry PRS methods will require substantially larger ancestry-matched discovery GWAS, underscoring the urgent need to expand genetic studies in underrepresented populations.

## INTRODUCTION

Parkinson’s Disease (PD) is a progressive neurodegenerative disorder primarily affecting movement. While 5-10% of PD cases are caused by monogenic inheritance, the vast majority are sporadic. Recent genome-wide association studies (GWAS) have uncovered dozens of loci that collectively explain up to ∼36% of risk in sporadic cases ^1–7^. Polygenic risk scores (PRS) aggregate the effects of multiple risk alleles into a single measure of genetic liability ^8^ and, in research settings, have shown promise in predicting PD status, particularly when combined with clinical ^9^ and neuroimaging ^10^ biomarkers. Likewise, integrating PRS with prodromal indicators, such as olfactory dysfunction, REM sleep behavior disorder, or dopaminergic imaging, can enhance early detection and improve discrimination between prodromal and diagnosed PD cases ^9^.

The predictive performance of PRS is highly dependent on the match between the ancestry of the discovery GWAS (from which SNP effect estimates are derived) and the ancestry of the target population in which those effects are aggregated into a PRS ^11–13^. Scores derived from predominantly European discovery GWAS often show reduced accuracy in non-European target populations ^14,15^, largely due to differences in allele frequencies and LD structure. Given that most large GWAS to date have been conducted in European samples,^16^ this imbalance raises concerns, as current PRS approaches risk exacerbating health disparities ^17^.

Latin American populations result from complex admixture among Native American, European, and African ancestral groups ^18,19^. This admixture has produced wide variation in global ancestry proportions between Latin American individuals, as well as substantial within individual variation in local ancestry proportions across the genome ^20,21^, rendering the portability of PRS especially challenging. In Latin America, PD is among the most prevalent neurodegenerative disorders, diagnosed in ∼2% in people older than 65 ^22^. The first GWAS of PD in Latin Americans (published in 2020) included 807 PD cases and 690 controls from five South American countries and identified known and novel genetic loci associated with PD ^4,23^. The Global Parkinson’s Genetics Program (GP2 ^24^) is an international initiative designed to expand the diversity and scale of PD genetic research, with a particular emphasis on underrepresented populations. GP2 builds on earlier GWAS efforts in Latin America by substantially increasing sample sizes, improving representation across the region, and enabling more powerful multi-ancestry analyses of PD risk and genetic architecture.

The suboptimal portability of PRS across ancestries has motivated the development of many PRS methods ^25–30^, broadly grouped into single- and multi-ancestry methods, depending on the number of discovery populations used in PGS construction. We focus on two single-ancestry methods, PRSice-2 ^31^, and SBayesRC ^32^, and two mutli-ancestry methods, PRS-CSx ^33^, and BridgePRS ^34^ because the methods are widely used, conceptually representative of major methodological classes, and offer a useful contrast between single-ancestry and multi-ancestry approaches to polygenic prediction.

PRSice-2 ^31^ implements the original approach to PRS construction, clumping and thresholding, which involves clumping variants based on LD, and retaining only marginally significant SNPs. While straightforward computationally efficient, the approach suffers from reduced accuracy when the discovery and target population ancestries are misaligned ^17^. SBayesRC ^32^ is a newer single-ancestry method that improves cross-ancestry portability using a mixture-of-normals prior that incorporates functional annotations, which are more consistent across populations.

Multi-ancestry methods incorporate effect sizes and LD information from multiple discovery populations of different ancestries. PRS-CSx ^33^ extends the Bayesian continuous shrinkage framework of PRS-CS to jointly model ancestry-specific summary statistics and LD reference panels, enabling adaptive borrowing of information across ancestries while retaining ancestry-specific architecture. PRS-CSx typically requires target sample tuning and validation datasets to weight ancestry-specific scores optimally. BridgePRS ^34^ takes a complementary approach by combining ancestry-specific parameter estimates through a hierarchical model that modulates the degree of information sharing across populations, enabling more robust performance in the presence of ancestry-specific LD differences. In this way, the method “bridges” information from a large source ancestry (e.g., European) to a smaller target ancestry (e.g., Latin American). BridgePRS also requires tuning and validation datasets to estimate posterior weights.

While PGS benchmarking studies are increasingly common ^25–30,35^, several aspects of current evaluations make it difficult to determine which PGS method is most appropriate for constructing PRS for PD in Latin American populations. Most benchmarking studies focus on relatively homogenous continental ancestry groups, with more limited evaluation in three-way admixed populations. Such populations present distinct methodological challenges because multiple ancestral LD structures and allele frequency distributions co-occur within individuals. Compared to two-way admixture, the presence of three ancestral components increases genomic heterogeneity and may amplify mismatches between discovery GWAS LD patterns and local ancestry–specific LD in the target sample. In addition, while multi-ancestry methods leverage the growing number of GWAS in non-European populations, the available sample size in practice often remains substantially smaller (e.g., 807 PD cases and 690 controls ^4^) than those used in method development and benchmarking studies (e.g., ∼5k individuals). These differences may affect real-world performance.

Our objective was to evaluate the accuracy of single- and multi-ancestry PRS methods in a sample of Latin American individuals with and without PD genotyped under GP2 efforts, using existing PD GWAS summary statistics. Results inform future genomic studies of PD in Latin American populations.

## MATERIALS & METHODS

### Target data

GP2 comprises over 300 participating studies worldwide, each with diverse recruitment strategies that adapt to regional capacities and medical systems; nevertheless, all PD cases must meet the UKBB/MDS criteria, controls have no neurological disease, and participating studies must submit at least minimal clinical data on participants ^36^.

We used GP2 release 9 genotyping data. Sample processing was standardised and harmonised by the GP2 consortium. Details on genotyping and quality control (QC) procedures are described elsewhere ^37^. Briefly, all samples were genotyped using the Illumina NeuroBooster array, an extension of the Infinium Global Diversity Array. This platform includes an additional 95,273 variants associated with neurological conditions, supplementing the 1,914,934 variants initially present in the Global Diversity Array ^38^. Following genotyping, all samples underwent imputation using the TOPmed imputation server^38^. Ancestry was assigned to each sample using Uniform Manifold Approximation and Projection (UMAP)^39^ and Extreme Gradient Boosting (XGBoost) ^40^, with reference populations from the 1000 Genomes Project ^41^ and the Human Genome Diversity Project ^42^. Ancestry-specific QC was then conducted separately for each group, including assessments of call rate, kinship, sex concordance, and heterozygosity. Additionally, variant-level QC included filtering based on haplotype missingness and Hardy-Weinberg equilibrium.

For this study, we selected samples labelled as “Latino and Indigenous people of the Americas” (i.e., those who mapped to the Admixed American (AMR) population in the 1000 Genomes Project), which included 3,395 individuals (1,945 PD-affected and 1,450 controls) with data on 65,288,233 single-nucleotide polymorphisms (SNPs). The geographic origin and participating studies from which these samples originate are shown in Supplementary Tables 1 and 2.

### Target data processing

To prepare our genotype data for PRS construction, we applied additional filters beyond the standard QC and harmonisation performed by GP2. We retained only well-imputed variants (imputation quality R² > 0.3) and performed all subsequent filtering using PLINK v2 ^43^. We converted genotype dosages to hard calls, applying a hard-call threshold of 0.1 and excluding indels, multiallelic variants, variants with a minor allele frequency (MAF) < 1%, or with > 5% missingness (--geno 0.05). Variant IDs were updated from Chromosome-Position-Ref-Alt format to rsIDs, using the dbSNP GRCh38 reference panel, and we removed unmapped and duplicated rsID SNPs. Individuals with > 2% missing genotypes were excluded (--mind 0.02). To account for potential cryptic relatedness, we used precomputed identity-by-descent (IBD) proportions provided by GP2 ^37,38^ and excluded one individual from each pair with an IBD proportion greater than 0.2. We additionally excluded individuals with an “unclear” PD diagnosis, as well as those with suspected monogenic forms of PD. After applying all variant and sample QC filters, 51,081,618 SNPs and 3,315 individuals remained.

To account for latent population stratification within our sample, we performed principal component analysis (PCA) on the 3,315 individuals. For this, genotyped SNPs only were LD-pruned in PLINK (--indep-pairwise 200 50 0.2), removing one variant from each pair with r² > 0.2 within sliding windows of 200 SNPs shifted by 50. Subsequently, we applied smartPCA ^44^ to the pruned dataset (98,373 SNPs) using the default parameters.

### Target data global ancestry

We estimated global ancestry proportions for all individuals in the target sample using ADMIXTURE (v1.3.0). We defined K = 3 and referenced African (AFR), Admixed American (AMR), and European (EUR) continental populations from the 1000 Genomes Project ^45^.

### PD GWAS summary statistics

We used three PD GWAS as our discovery sample datasets (Figure 1): (i) The EUR GWAS summary statistics were computed from the largest PD GWAS meta-analysis to date, comprising 63,555 PD cases, 17,700 proxy cases (individuals with a family history of PD), and 1,746,386 controls^46^. (ii) The AMR GWAS summary statistics originated from the most extensive Latin American PD GWAS, which included 807 PD cases and 690 controls^4^. (iii) The multi-ancestry meta-analysis (MAMA) GWAS summary statistics included individuals from EUR, AMR, East Asian (EAS), and African (AFR) populations, encompassing 49,049 cases, 18,785 proxy cases, and 2,458,063 controls ^5^.

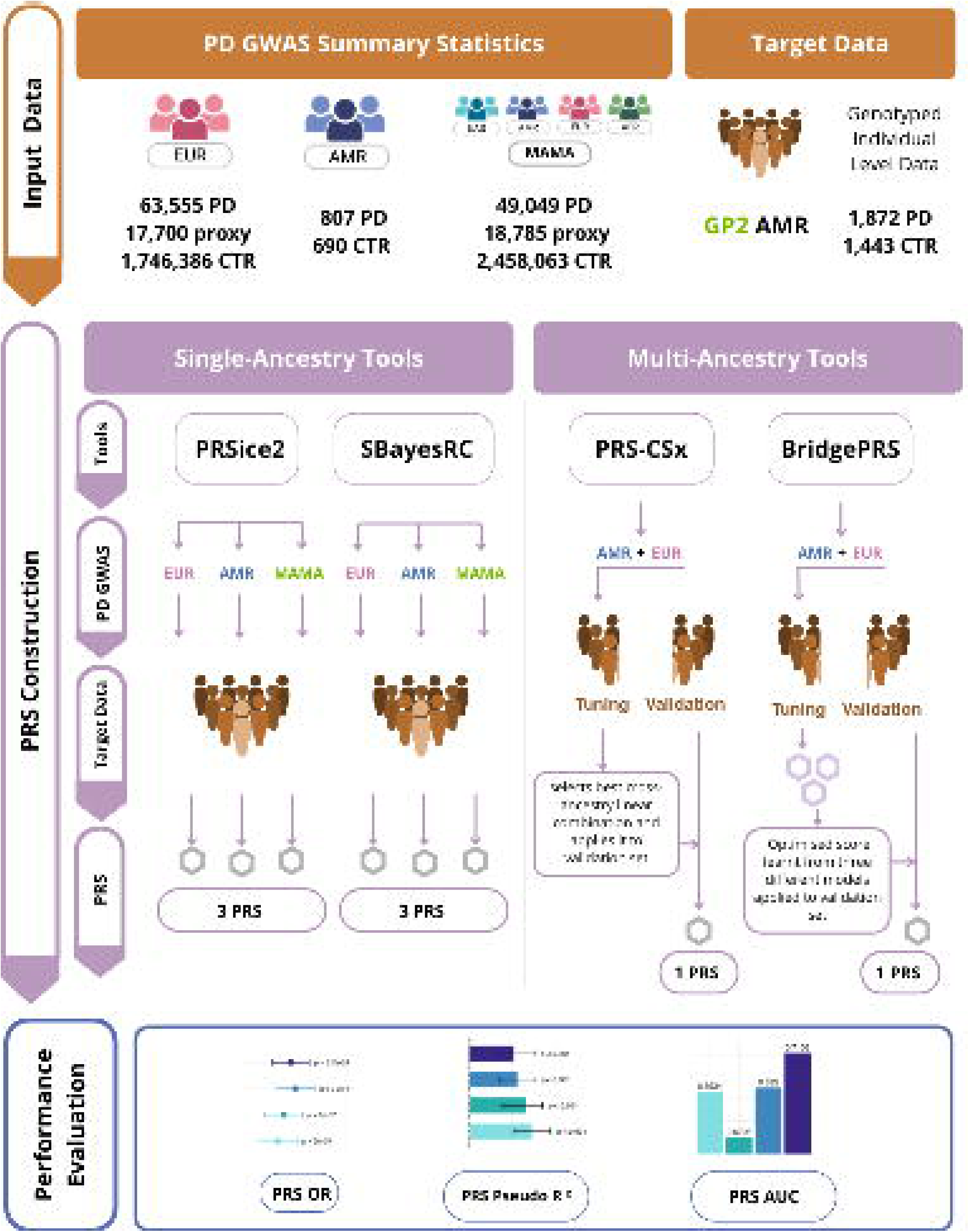

To ensure that all summary statistics were in a standardised format required for PRS development, we used the MungeSumstats R package (V 1.12.2) with default parameters ^47^. The MungeSumstats workflow aligned alleles and variant positions to the reference genome build (GRCh38), removed strand-ambiguous SNPs, and ensured the effect allele was consistent with the reference. It verified key fields (rs IDs, chromosome, position, alleles, effect sizes, standard errors, and P-values), and ensured values fell within plausible ranges (MAF > 0.05). Following QC, 10,372,428 variants remained in the EUR summary statistics, 4,443,812 in the AMR summary statistics, and 7,206,410 in the MAMA summary statistics.

### PRS construction

We selected four tools to compute PRS in our target population: two single-ancestry methods (PRSice-2 and SBayesRC) and two multi-ancestry methods (PRS-CSx and BridgePRS). Our general workflow is presented in Figure 1. Since the single-ancestry methods (PRSice-2 and SBayesRC) only allow a single discovery sample as input, we calculated PRS using summary statistics from each of the EUR, AMR, and MAMA datasets separately, producing three PRS for each method. For multi-ancestry methods, PRS-CSx and BridgePRS, we input both the EUR and AMR summary statistics simultaneously and calculated a single PRS. To enable hyperparameter tuning in the multi-ancestry methods, we randomly split the target dataset into a tuning subset (983 cases and 675 controls) and a validation subset (889 cases and 768 controls).

The methodological details of these tools have been described elsewhere ^31–33^. We briefly summarize them here for clarity and provide additional implementation details in Supplementary Tables 3 and 4.

PRSice-2 employs a Clumping and Thresholding strategy. It prunes variants in LD to keep independent variants and tests multiple p-value thresholds to progressively include more variants in the PRS model ^31^. We employed this tool using its default clumping-and-thresholding approach (r² = 0.1, 250 kb window) across eight p-value thresholds (P ≤ 0.001, 0.05, 0.1, 0.2, 0.3, 0.4, 0.5, and 1) and selected the threshold that maximized Nagelkerke’s pseudo R^2^ of the PRS (Supplementary Table 3) ^31^.

We applied SBayesRC using default program inputs and parameters. Posterior SNP effect sizes generated by SBayesRC were used to compute PRS in the target dataset without the need for additional tuning^32^.

PRS-CSx calculates ancestry-specific posterior SNP effect sizes using a Bayesian regression framework with coupled global-local continuous shrinkage (CS) priors, where the global parameter (Φ) controls the overall sparsity of the genetic architecture and the local parameters allow individual SNPs with strong evidence to escape shrinkage. During this step, ancestry-matched LD reference panels are incorporated to model population-specific correlation structure among SNPs. The model then constructs ancestry-specific PRS and learns an optimal linear combination of these scores in a validation set from the target population. In this tuning step, the global shrinkage parameter and the combination weights are optimised. The resulting weighted PRS is then applied to an independent validation dataset to evaluate predictive performance^33^. We computed PRS using four values of Φ (1, 1e−2, 1e−4, and 1e−6), generating ancestry-specific SNP-weight files for EUR and AMR for each value of Φ. We then used PLINK v2 (--score) to compute PRS values for each ancestry-specific score. Next, we manually evaluated all linear combinations of varying Φ EUR and AMR scores on the tuning subset. The linear combination yielding the highest pseudo-R² value for case–control status was selected (Φ = 1e−4 for both EUR and AMR, Supplementary Table 4). This best-performing linear combination was then carried forward for use in the validation subset ^33^.

BridgePRS computes three models based on different assumptions about the informativeness of GWAS data across populations. Model 1 applies a zero-centred Gaussian prior to SNP effect sizes in the discovery GWAS (EUR) and then uses the resulting estimates as priors for the target population (AMR), assuming the EUR GWAS is the most informative. Model 2 applies Model 1 zero-centred Gaussian prior analysis directly to the target population alone, assuming it is the only informative source. Model 3 combines the PRS from Models 1 and 2 using ridge regression in the target population, considering that both GWAS provide independent information. Since the true architecture is unknown, BridgePRS averages predictions across all three models ^34^. We tuned the shrinkage parameter governing the extent of effect-size sharing across ancestries in the tuning subset, and the optimized score was evaluated in the validation subset using PLINK v2.

#### Evaluation of PRS predictive performance

We evaluated the accuracy of the PRS using three complementary metrics within a logistic regression framework: (i) odds ratios (OR) to quantify the relative change in PD risk per SD increase in PRS; (ii) Nagelkerke’s pseudo R^2^ to quantify the proportion of variance in PD risk explained by the PRS; and (iii) area under the receiver operating characteristic curve (AUC), assessing the discriminative ability of the PRS to distinguish cases from controls.

All logistic regression models were implemented in R v4.4.2 (glm package) and included sex, age, the first 10 principal components (PCs), and family history as covariates. Individuals with missing age information were excluded from the regression. Family history was coded as a categorical variable with three levels: positive, negative, and unknown. We computed Nagelkerke’s pseudo R² using the pseudo-R² function from the DescTools (R v4.3.1) package (v0.99.60). Values were transformed to a liability scale using the following formula:

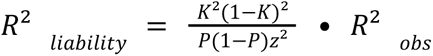

where K is the population prevalence of PD (0.5%), P is the simple case fraction, and z is the height of the normal density at the threshold for K ^48^. We estimated 95% confidence intervals (CIs) for each pseudo-R² via 2,000 bootstrap iterations, using the boot.ci function from the boot (R v4.3.1) package (v1.3.31) with the percentile (perc) method. To estimate PRS-specific discrimination independent of covariates, we first fitted a covariate-only logistic regression model and extracted the residuals. We then regressed these residuals on the standardized PRS and used the resulting predicted values to construct receiver operating characteristic curves against case–control status and the corresponding PRS-specific AUC. Analyses were completed using the pROC ( R v4.3.1) package (v1.19.0.1).

### Predictive performance by ancestry composition

Ultimately, PRS constructed using SBayesRC and EUR summary statistics explained the most variance in PD risk (the highest liability-scale pseudo-R²). We next investigated the influence of ancestry composition on PRS performance using this tool only. We stratified the cohort into four EUR ancestry quartiles based on individual-level EUR ancestry proportions (cutoffs for EUR% = 0.0000–0.1464, 0.1464–0.4378, 0.4378–0.6268, and 0.6268–1.0000). To ensure balanced group comparisons, we identified the maximum possible number of cases and controls across quartiles and downsampled all groups accordingly (197 cases and 173 controls). Principal components were recalculated within each quartile using PLINK v2 (--pca). Within each group, we evaluated PRS performance as above.

## RESULTS

After QC, the final GP2 AMR target dataset comprised 1,872 PD cases and 1,443 controls originating from 17 different countries and participating studies (Supplementary Tables 1 and 2). Cases were older than controls and were less likely to be female (Table 1). Cases were also more likely to have a family history of PD, although family history was missing in 34% of cases and 12% of controls. The admixture proportions of cases and controls also differed: cases had a greater proportion of EUR and AFR ancestry, while controls had a greater proportion of AMR ancestry. These observed differences in demography (age, sex), clinical risk factors (family history), and global ancestry proportions between cases and controls highlight the need to include these factors (age, sex, and the first 10 PCs) as covariates in all subsequent logistic regression models.

**Table 1.** Characteristics of GP2 participants of AMR ancestry included in this study after QC.

|  | PD Cases | Controls |
| --- | --- | --- |
| **N** | 1872 | 1443 |
| **Age in years, mean (SD)** | 63.5 (12.1) | 59.94 (8.5) |
| Missing age, N (%) | 67 (3.6%) | 53 (3.7%) |
| **Age at PD onset in years, mean (SD)** | 56.77 (12.8) | — |
| Missing age at PD onset, N (%) | 1556 (83.1%) | — |
| **Female sex, N (%)** | 818 (43.7%) | 935 (64.8%) |
| **Family history of PD** |  |  |
| Yes, N (%) | 308 (16.5%) | 80 (5.5%) |
| No, N (%) | 919 (49.1%) | 1185 (82.1%) |
| Missing family history, N (%) | 645 (34.5%) | 178 (12.3%) |
| **Global genetic ancestry proportions** |  |  |
| EUR, mean (SD) | 0.505 (0.235) | 0.290 (0.264) |
| AFR, mean (SD) | 0.043 (0.054) | 0.021 (0.040) |
| AMR, mean (SD) | 0.452 (0.249) | 0.688 (0.281) |

No single combination of PRS method and GWAS discovery sample performed the best across all three metrics of predictive performance (Figure 2; Supplementary Table 5). A PRS constructed using SBayesRC and the EUR GWAS achieved the highest explanatory power, outranking other methods in terms of both OR and liability-scale pseudo-R². A PRS constructed using SBayesRC and the MAMA GWAS had the highest discriminatory power, as determined by the AUC. PRS-CSx constructed with EUR + AMR summary statistics also performed well across all metrics, consistently outranking PRSice2 and BridgePRS.

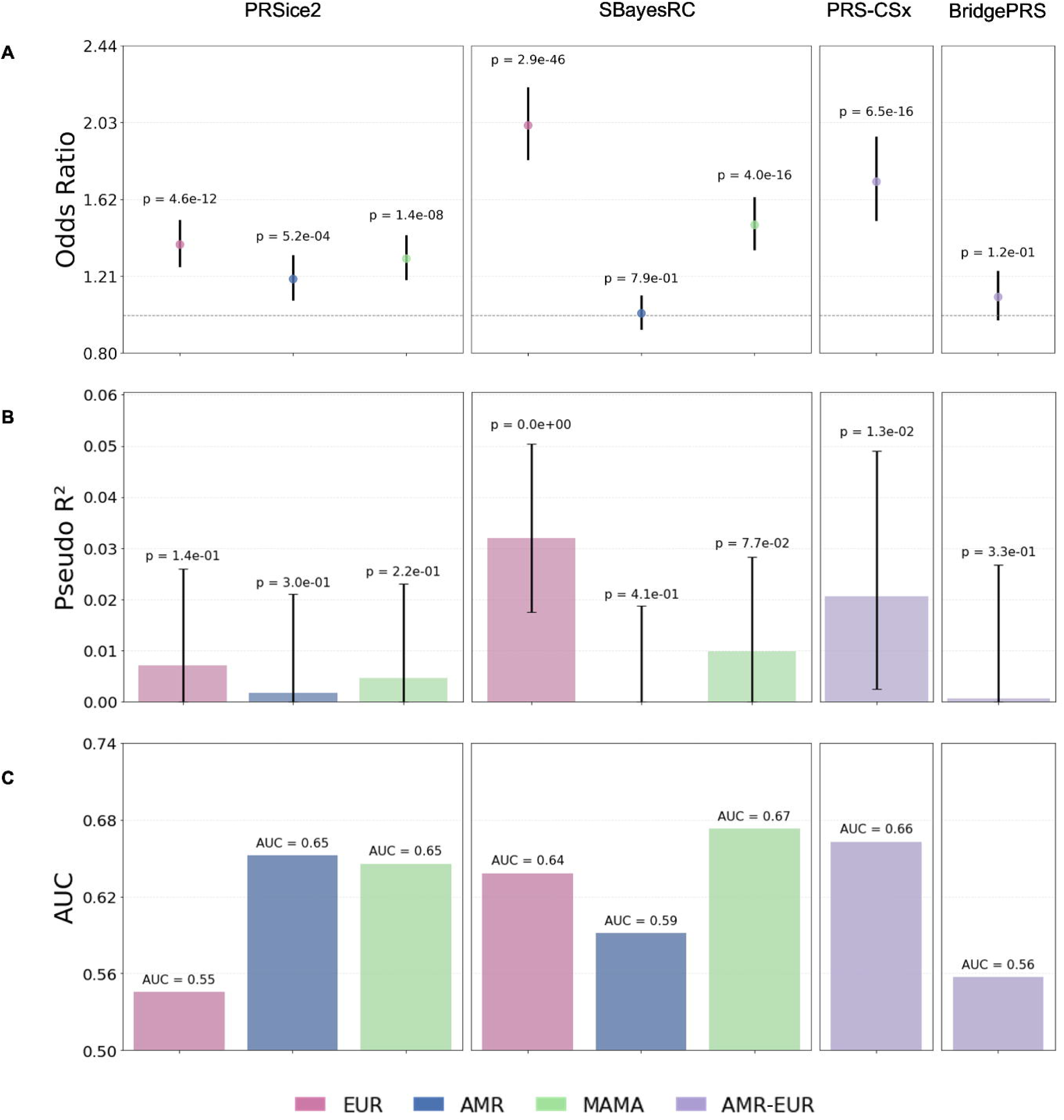

We next assessed how differences in the proportion of global EUR ancestry between individuals contributed to the PRS predictive performance. We stratified our target sample by EUR ancestry quartiles, and focused on the PRS that performed well across multiple metrics in the full sample, i.e., the PRS constructed using SBayesRC and the EUR GWAS. The PRS was significantly associated with PD status in all quartiles, with ORs ranging from 1.85 in Q1 (lowest EUR ancestry) (p = 2.0×10⁻⁵) to 2.40 in Q3 (p = 9.2×10⁻¹⁰), indicating that each standard deviation increase in PRS was associated with approximately a two-fold increase in the odds of Parkinson’s disease. The liability-scale PRS pseudo R^2^ varied across quartiles, from 0.047 in Q1 to 0.1 in Q2, while discrimination remained stable, with AUC values ranging from 0.67 in Q1 to 0.71 in Q4 (Figure 3; Supplementary Table 6).

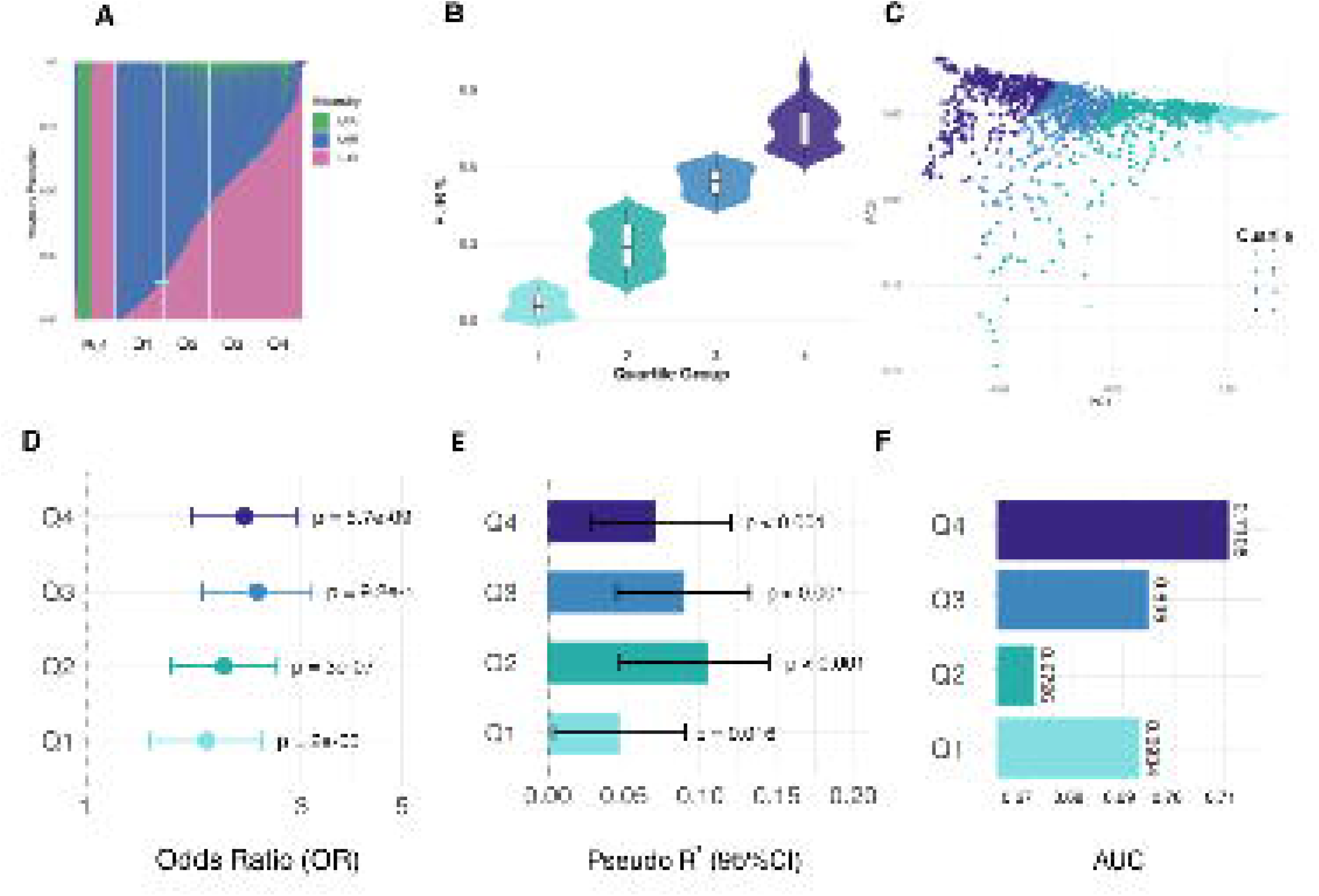

Finally, we sought to contextualize the predictive performance of a PRS for PD alongside conventional clinical PD risk factors. To do this, we evaluated a series of nested logistic regression models. These models were (i) a basic model including age and sex; (ii) an intermediate model including age, sex, and family history of PD; and (iii) a full model including age, sex, family history, and a PRS constructed using SBayesRC and the EUR GWAS. Genetic PCs were not included in this model because the purpose was to quantify the discriminatory ability of the PRS itself beyond clinical risk factors, rather than the combined predictive contribution of ancestry and the PRS. This follows the rationale in other PRS benchmarking studies ^49^. As variables were added to each model, discrimination and proportion of phenotypic variance explained improved (Figure 4A). The full model achieved the highest discriminative accuracy (AUC = 0.728) and explained the largest proportion of variance (Nagelkerke R² = 0.2). The polygenic risk score was strongly associated with disease risk (OR = 2.02, 95% CI = 1.83 – 2.22, P < 2.94×10⁻46), independent of age, sex, family history, and genotype PCs (Figure 4B; Supplementary Table 7).

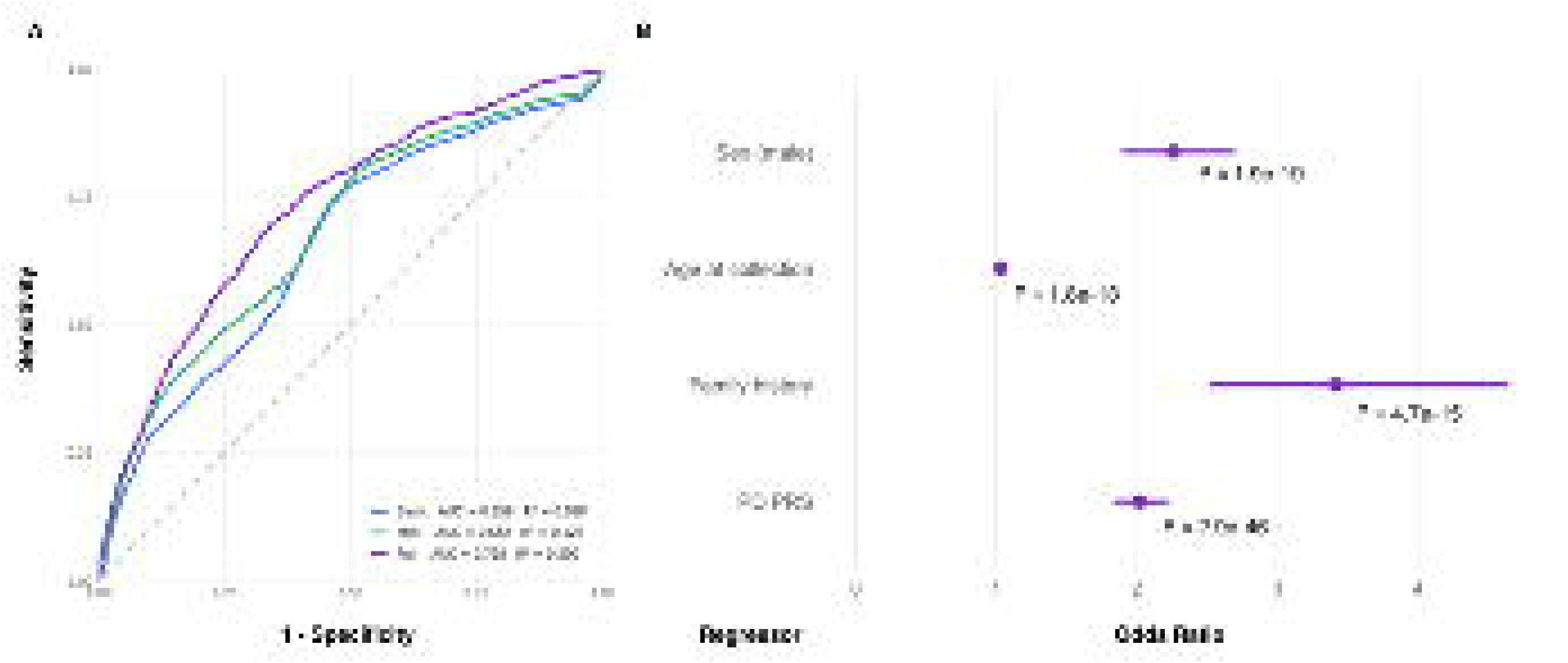

## DISCUSSION

Our PRS benchmarking study in a Latin American sample of PD cases and controls found that PRS predictive ability depended on the PRS tool, discovery GWAS summary statistics, and performance measure. The single-ancestry method SBayesRC using EUR GWAS summary statistics achieved the strongest risk estimates (OR) and highest explanatory power (pseudo-R²), whereas SBayesRC using MAMA GWAS summary statistics had the best discriminative ability (AUC). Stratification by quartiles of global European ancestry showed that predictive performance was higher in the top vs. bottom quartile groups, consistent with improved performance when the target population is genetically closer to the discovery sample ^35,50,51^. Including a PRS alongside conventional sociodemographic and clinical predictors showed that despite modest predictive ability of the PRS in absolute terms, the OR of the PRS was comparable to other risk factors in the model, and improved model performance as measured by the AUC.

Our results reflect the current reality in polygenic risk prediction for underrepresented populations: ancestry-matched GWAS data may exist, but are often severely underpowered relative to EUR GWAS, limiting their predictive utility ^52^. The EUR dataset comprised over 80,000 PD cases and proxy cases, compared with fewer than 1,000 AMR cases. Prior work has shown that when ancestry-diverse discovery GWAS are better powered, multi-ancestry tools can match or outperform single-ancestry tools using large European-only GWAS summary statistics ^53^, suggesting that the underperformance of multi-ancestry tools here reflects limited AMR discovery sample size rather than a fundamental shortcoming of the methods. [Pivot to the need for more diverse GWAS, which GP2 is addressing. These methods therefore warrant ongoing investigation as sample sizes of diverse GWAS increase.

Larger discovery sample sizes, as seen in the SBayesRC EUR analyses, appear to primarily improve the proportion of PD liability explained by genetic risk, whereas incorporating multi-ancestry effect estimates (MAMA GWAS) improved the discriminatory performance. These results are intuitive, as greater power (larger GWAS) allows more risk variants to be mapped, increasing the variance explained. In contrast, the discriminatory potential of SBayesRC with MAMA GWAS summary statistics likely reflects the complementary advantages of leveraging both functional genomic annotations and diverse GWAS data. Multi-ancestry meta-analyses improve the estimation of effect sizes across varied linkage disequilibrium (LD) and allele frequency backgrounds, thereby enhancing portability to admixed populations. At the same time, SBayesRC incorporates functional annotations that enrich for variants more likely to represent true causal effects, which are often substantially shared across ancestries despite differences in LD structure ^54–57^. By jointly modeling cross-population effect size information and biologically informed priors, SBayesRC may preferentially capture components of genetic architecture that are robust to ancestry heterogeneity. As multi-ancestry GWAS continue to expand, methods that explicitly integrate both genetic diversity and functional biology may be particularly well positioned to improve polygenic risk prediction in admixed populations. Defining the functional variants (computationally or experimentally) may also improve power and transferability of PRS across populations.

When stratifying the cohort by quartiles of global European ancestry, we observed a discordant pattern whereby Nagelkerke’s pseudo-R² was highest in the second quartile, while the AUC was lowest. Relative to the other quartiles, the second quartile was most heterogenous with regards to global ancestry proportions, as evidenced by the wider dispersion along PC1 and PC2 (Figure 3c). Therefore, the discordant pattern comparing R² and AUC values may reflect increased variability in PRS values in the second quartile, which can increase likelihood-based measures of variance explained without a corresponding improvement in case-control discrimination. In contrast to the AUC, which depends solely on rank ordering, pseudo-R² metrics are sensitive to the distribution and scale of the predictor. This observation underscores the sensitivity of performance metrics to ancestry heterogeneity within admixed populations, a consideration that is especially relevant to Latin American populations, which are exceptionally diverse, encompassing millions of people across multiple countries with complex demographic histories and varied environments.

Our study has several limitations. First, although most individuals were recruited in Latin America, the “AMR” label was assigned post hoc using global ancestry estimation and clustering, and some participants may have originated from non–Latin American countries. Second, we used a case-control analysis, and the liability-scale transformation assumes a uniform PD prevalence, which may not hold across subpopulations. Third, the EUR and MAMA GWAS summary statistics incorporated proxy cases (individuals reporting a family history of PD). While such proxy phenotypes increase statistical power, they may also introduce biases related to survival, reporting accuracy, and non-random participation, as highlighted in studies of other late-onset neurodegenerative disorders ^58^. While these biases could attenuate polygenic risk predictions, comparisons across methods using the same summary statistics, as we have done, are still valid. Fourth, we did not evaluate model calibration, a measure of concordance between predicted and observed risk ^59^, which is an essential precursor to absolute risk estimates and clinical implementation. Methods for calibration in diverse ancestry samples is an active area of research ^60,61^. Fifth, we assigned individuals to discrete genetic ancestry groups and evaluated predictive performance at the population level, an approach commonly used in PRS evaluations given the structure of available GWAS data. However, this strategy overlooks that genetic ancestry lies on a continuum and that the accuracy of polygenic scores can vary continuously across that ancestry spectrum, even within traditionally defined groups ^11^. As genotype data in diverse populations continues to grow, future work should evaluate PRS performance at the level of individual genetic ancestry, which may provide a more nuanced understanding of prediction accuracy and portability in admixed samples.

In conclusion, our study demonstrates that in an admixed Latin American sample, a PD PRS derived from a large EUR GWAS outperforms ancestry-matched but underpowered discovery datasets, with predictive performance increasing alongside European ancestry proportion. While current multi-ancestry approaches provide incremental benefits, their advantages will likely grow as GWAS in diverse populations increase in size and representation. Together, these findings underscore both the potential and the challenges of equitable PRS implementation in genetically diverse populations, as well as the need for larger, ancestrally inclusive genetic discovery efforts.

## Data Availability

All data produced in the present work are contained in the manuscript

https://www.amp-pd.org

https://www.cog-genomics.org/plink/2.0/resources

## Acknowledgements

V.F.-O. is a recipient of a scholarship from the GP2 Trainee Network, part of the Global Parkinson’s Genetics Program, and funded by the Aligning Science Across Parkinson’s (ASAP) initiative. M.E.R. thanks the support from the Rebecca L. Cooper Medical Research Foundation (F20231230) and a Pilot Award for Global Brain Health Leaders by the Global Brain Health Institute, Alzheimer’s Association, and Alzheimer’s Society (GBHI ALZ UK-22-869020). JKD is a Michael Smith Health Research BC Scholar and her work is supported by NSERC (RGPIN-2024-06087).

This project was supported by the Global Parkinson’s Genetics Program (GP2; https://gp2.org). GP2 is funded by the Aligning Science Across Parkinson’s (ASAP) initiative and implemented by The Michael J. Fox Foundation for Parkinson’s Research (MJFF). For a complete list of GP2 members, see https://doi.org/10.5281/zenodo.7904831.

## Author Contributions

V.F.O.: study design, data analysis, and manuscript writing.

P.R.P.: analyses support, manuscript writing, and figure preparation/visualization.

N.S.O.: analyses support and manuscript writing.

G.S.-P.: figure preparation/visualization and manuscript revision.

S.D.-T.: analyses support and Discussion drafting.

P.A.P.: project concept and study outline.

T.P.L., E.W., A.E.R.C., and S.A.: methodological input.

I.F.M. and M.E.R.: methodological guidance.

A.M.R. and J.K.D.: supervision and project outline.

GP2.: provision of data resources and computational infrastructure support.

All authors: manuscript editing and final approval.

## Data Availability

Data used in the preparation of this article were obtained from the Global Parkinson’s Genetics Program (GP2; https://gp2.org). Specifically, we used Tier 2 data from GP2 release 9 (https://doi.org/10.5281/zenodo.14510099). Access to the GP2 data can be obtained through an application via the AMP-PD platform (https://amp-pd.org). The One Thousand Genomes dataset was used for Global Ancestry estimation and can be accessed in Plink format via the PLINK resource platform (https://www.cog-genomics.org/plink/2.0/resources)

## Code Availability

All code generated for this article, and the identifiers for all software programs and packages used, are available on GitHub (https://github.com/GP2code/Cross_ancestry_PRS_AMR/tree/main) and were given a persistent identifier via Zenodo (DOI: https://doi.org/10.5281/zenodo.18831467).

## REFERENCES

1. Blauwendraat, C., Nalls, M. A. & Singleton, A. B. The genetic architecture of Parkinson’s disease. Lancet Neurol. 19, 170–178 (2020).

2. Cherian, A. & Divya, K. P. Genetics of Parkinson’s disease. Acta Neurol. Belg. 120, 1297–1305 (2020).

3. Nalls, M. A. et al. Identification of novel risk loci, causal insights, and heritable risk for Parkinson’s disease: a meta-analysis of genome-wide association studies. Lancet Neurol. 18, (2019).

4. Loesch, D. P. et al. Characterizing the Genetic Architecture of Parkinson’s Disease in Latinos. Ann. Neurol. 90, (2021).

5. Kim, J. J. et al. Multi-ancestry genome-wide association meta-analysis of Parkinson’s disease. Nat. Genet. 56, 27–36 (2024).

6. Rizig, M. et al. Identification of genetic risk loci and causal insights associated with Parkinson’s disease in African and African admixed populations: a genome-wide association study. Lancet Neurol. 22, 1015–1025 (2023).

7. Foo, J. N. et al. Genome-wide association study of Parkinson’s disease in East Asians. Hum. Mol. Genet. ddw379 (2016).

8. Lewis, C. M. & Vassos, E. Polygenic risk scores: from research tools to clinical instruments. Genome Med. 12, 1–11 (2020).

9. Maraki, M. I. et al. Association of the polygenic risk score with the probability of prodromal Parkinson’s disease in older adults. Front. Mol. Neurosci. 14, 739571 (2021).

10. Kals, M. et al. Polygenic risk score combined with transcranial sonography refines Parkinson’s disease risk prediction. Mov. Disord. Clin. Pract. 12, 928–937 (2025).

11. Ding, Y. et al. Polygenic scoring accuracy varies across the genetic ancestry continuum. Nature 618, 774–781 (2023).

12. Kachuri, L. et al. Principles and methods for transferring polygenic risk scores across global populations. Nat. Rev. Genet. 25, 8–25 (2024).

13. Wang, Y. et al. Theoretical and empirical quantification of the accuracy of polygenic scores in ancestry divergent populations. Nat. Commun. 11, 3865 (2020).

14. Martin, A. R. et al. Human Demographic History Impacts Genetic Risk Prediction across Diverse Populations. Am. J. Hum. Genet. 100, (2017).

15. Duncan, L. et al. Analysis of polygenic risk score usage and performance in diverse human populations. Nat. Commun. 10, 3328 (2019).

16. Peterson, R. E. et al. Genome-wide Association Studies in Ancestrally Diverse Populations: Opportunities, Methods, Pitfalls, and Recommendations. Cell 179, 589–603 (2019).

17. Martin, A. R. et al. Clinical use of current polygenic risk scores may exacerbate health disparities. Nat. Genet. 51, 584–591 (2019).

18. Ruiz-Linares, A. et al. Admixture in Latin America: geographic structure, phenotypic diversity and self-perception of ancestry based on 7,342 individuals. PLoS Genet 10, e1004572 (2014).

19. Price, A. L. et al. A genomewide admixture map for Latino populations. Am J Hum Genet 80, 1024–1036 (2007).

20. Norris, E. T. et al. Genetic ancestry, admixture and health determinants in Latin America. BMC Genomics 19, 861 (2018).

21. Mendoza-Revilla, J. et al. Disentangling signatures of selection before and after European colonization in Latin Americans. Mol. Biol. Evol. 39, (2022).

22. Prevalence of parkinsonism and Parkinson disease in urban and rural populations from Latin America: A community based study. The Lancet Regional Health - Americas 7, 100136 (2022).

23. Elsayed, I. & Foote, I. F. Improving diversity in Parkinson’s disease genetics: Findings from the first-ever genome-wide association study in Latinos. Mov. Disord. 36, 2505 (2021).

24. Global Parkinson’s Genetics Program. GP2: The global Parkinson’s genetics program. Mov. Disord. 36, 842–851 (2021).

25. Sun, Q. et al. Improving polygenic risk prediction in admixed populations by explicitly modeling ancestral-differential effects via GAUDI. Nat. Commun. 15, 1016 (2024).

26. Privé, F., Arbel, J. & Vilhjálmsson, B. J. LDpred2: better, faster, stronger. Bioinformatics 36, 5424–5431 (2021).

27. Albiñana, C. et al. Multi-PGS enhances polygenic prediction by combining 937 polygenic scores. Nat. Commun. 14, 4702 (2023).

28. Marnetto, D. et al. Ancestry deconvolution and partial polygenic score can improve susceptibility predictions in recently admixed individuals. Nat. Commun. 11, 1628 (2020).

29. Cai, M. et al. A unified framework for cross-population trait prediction by leveraging the genetic correlation of polygenic traits. Am. J. Hum. Genet. 108, 632–655 (2021).

30. Benoumhani, S. et al. A review of methods and software for polygenic risk score analysis. PeerJ Comput. Sci. 11, e3039 (2025).

31. Choi, S. W. & O’Reilly, P. F. PRSice-2: Polygenic Risk Score software for biobank-scale data. Gigascience 8, (2019).

32. Zheng, Z. et al. Leveraging functional genomic annotations and genome coverage to improve polygenic prediction of complex traits within and between ancestries. bioRxiv 2022.10.12.510418 (2022) doi:10.1101/2022.10.12.510418.

33. Ruan, Y. et al. Improving polygenic prediction in ancestrally diverse populations. Nat. Genet. 54, 573–580 (2022).

34. Hoggart, C. J. et al. BridgePRS leverages shared genetic effects across ancestries to increase polygenic risk score portability. Nat. Genet. 56, 180–186 (2024).

35. Saffie-Awad, P. et al. Insights into Ancestral Diversity in Parkinson’s Disease Risk: A Comparative Assessment of Polygenic Risk Scores. medRxiv 2023.11.28.23299090 (2024) doi:10.1101/2023.11.28.23299090.

36. Towns, C. et al. Defining the causes of sporadic Parkinson’s disease in the global Parkinson’s genetics program (GP2). NPJ Parkinsons Dis. 9, 131 (2023).

37. Vitale, D. et al. GenoTools: an open-source Python package for efficient genotype data quality control and analysis. G3 (Bethesda) 15, (2025).

38. Bandres-Ciga, S. et al. NeuroBooster Array: A Genome-Wide Genotyping Platform to Study Neurological Disorders Across Diverse Populations. medRxiv (2023) doi:10.1101/2023.11.06.23298176.

39. McInnes, L., Healy, J., Saul, N. & Großberger, L. UMAP: Uniform Manifold Approximation and Projection. J. Open Source Softw. 3, 861 (2018).

40. Chen, T. & Guestrin, C. XGBoost. in Proceedings of the 22nd ACM SIGKDD International Conference on Knowledge Discovery and Data Mining (ACM, New York, NY, USA, 2016). doi:10.1145/2939672.2939785.

41. Byrska-Bishop, M. et al. High-coverage whole-genome sequencing of the expanded 1000 Genomes Project cohort including 602 trios. Cell 185, 3426–3440.e19 (2022).

42. Bergström, A. et al. Insights into human genetic variation and population history from 929 diverse genomes. Science 367, eaay5012 (2020).

43. Chang, C. C. et al. Second-generation PLINK: rising to the challenge of larger and richer datasets. Gigascience 4, 7 (2015).

44. Smart Principal Component Analysis. https://christianhuber.github.io/smartsnp/reference/smart_pca.html.

45. 1000 Genomes Project Consortium et al. A global reference for human genetic variation. Nature 526, 68–74 (2015).

46. Leonard, H. L. Novel Parkinson’s Disease Genetic Risk Factors Within and Across European Populations. Genetic and Genomic Medicine (2025).

47. Murphy, A. E., Schilder, B. M. & Skene, N. G. MungeSumstats: a Bioconductor package for the standardization and quality control of many GWAS summary statistics. Bioinformatics 37, 4593–4596 (2021).

48. Lee, S. H., Goddard, M. E., Wray, N. R. & Visscher, P. M. A better coefficient of determination for genetic profile analysis: A better coefficient of determination. Genet. Epidemiol. 36, 214–224 (2012).

49. Ni, G. et al. A comparison of ten polygenic score methods for psychiatric disorders applied across multiple cohorts. Biol. Psychiatry 90, 611–620 (2021).

50. Wang, Y. et al. Theoretical and empirical quantification of the accuracy of polygenic scores in ancestry divergent populations. Nat. Commun. 11, (2020).

51. Bitarello, B. D. & Mathieson, I. Polygenic scores for height in admixed populations. G3 (Bethesda) 10, 4027–4036 (2020).

52. Wang, Y. et al. Global Biobank analyses provide lessons for developing polygenic risk scores across diverse cohorts. Cell Genom. 3, 100241 (2023).

53. Wang, Y., Tsuo, K., Kanai, M., Neale, B. M. & Martin, A. R. Challenges and opportunities for developing more generalizable polygenic risk scores. Annu. Rev. Biomed. Data Sci. 5, 293–320 (2022).

54. Amariuta, T. et al. Improving the trans-ancestry portability of polygenic risk scores by prioritizing variants in predicted cell-type-specific regulatory elements. Nat. Genet. 52, 1346–1354 (2020).

55. Weissbrod, O. et al. Leveraging fine-mapping and multipopulation training data to improve cross-population polygenic risk scores. Nat. Genet. 54, 450–458 (2022).

56. Weissbrod, O. et al. Functionally informed fine-mapping and polygenic localization of complex trait heritability. Nat. Genet. 52, 1355–1363 (2020).

57. Miao, J. et al. Quantifying portable genetic effects and improving cross-ancestry genetic prediction with GWAS summary statistics. Nat. Commun. 14, 832 (2023).

58. Wu, Y. et al. Pervasive biases in proxy genome-wide association studies based on parental history of Alzheimer’s disease. Nat. Genet. 56, 2696–2703 (2024).

59. Wand, H. et al. Improving reporting standards for polygenic scores in risk prediction studies. Nature 591, 211–219 (2021).

60. Hou, K. et al. Calibrated prediction intervals for polygenic scores across diverse contexts. Nat. Genet. 56, 1386–1396 (2024).

61. Lukas, E., Uffelmann, E., Treur, J. L. & Peyrot, W. 32. Estimating disorder probability in diverse ancestral backgrounds based on polygenic prediction using Bayesian polygenic score probability conversion x. Eur. Neuropsychopharmacol. 99, 69–70 (2025).

